## Supplementary figures and images for "COVIDHunter: An Accurate, Flexible, and Environment-Aware Open-Source COVID-19 Outbreak Simulation Model"

### PaperFigures.pptx

#### Slide 1
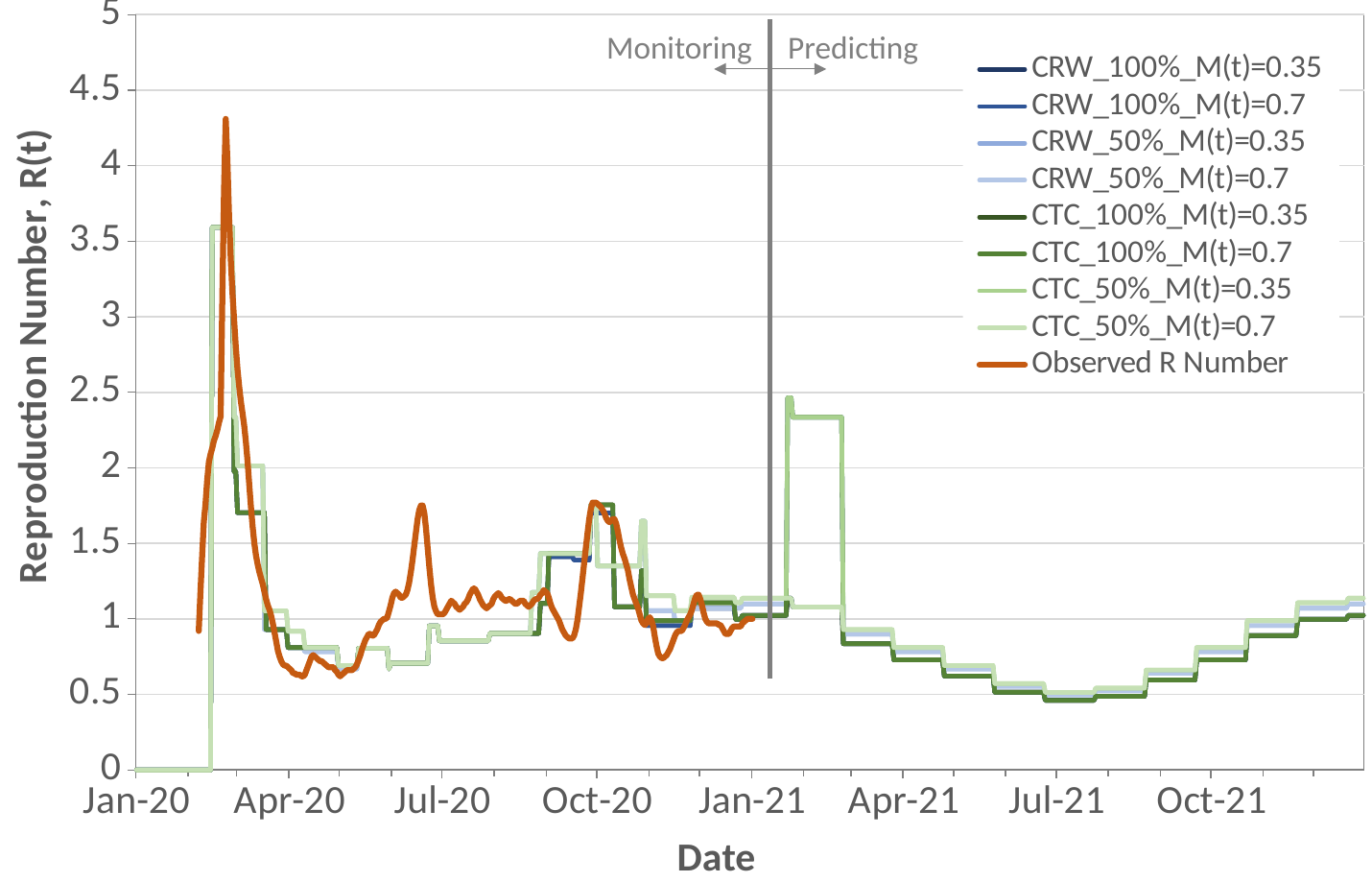

#### Slide 2
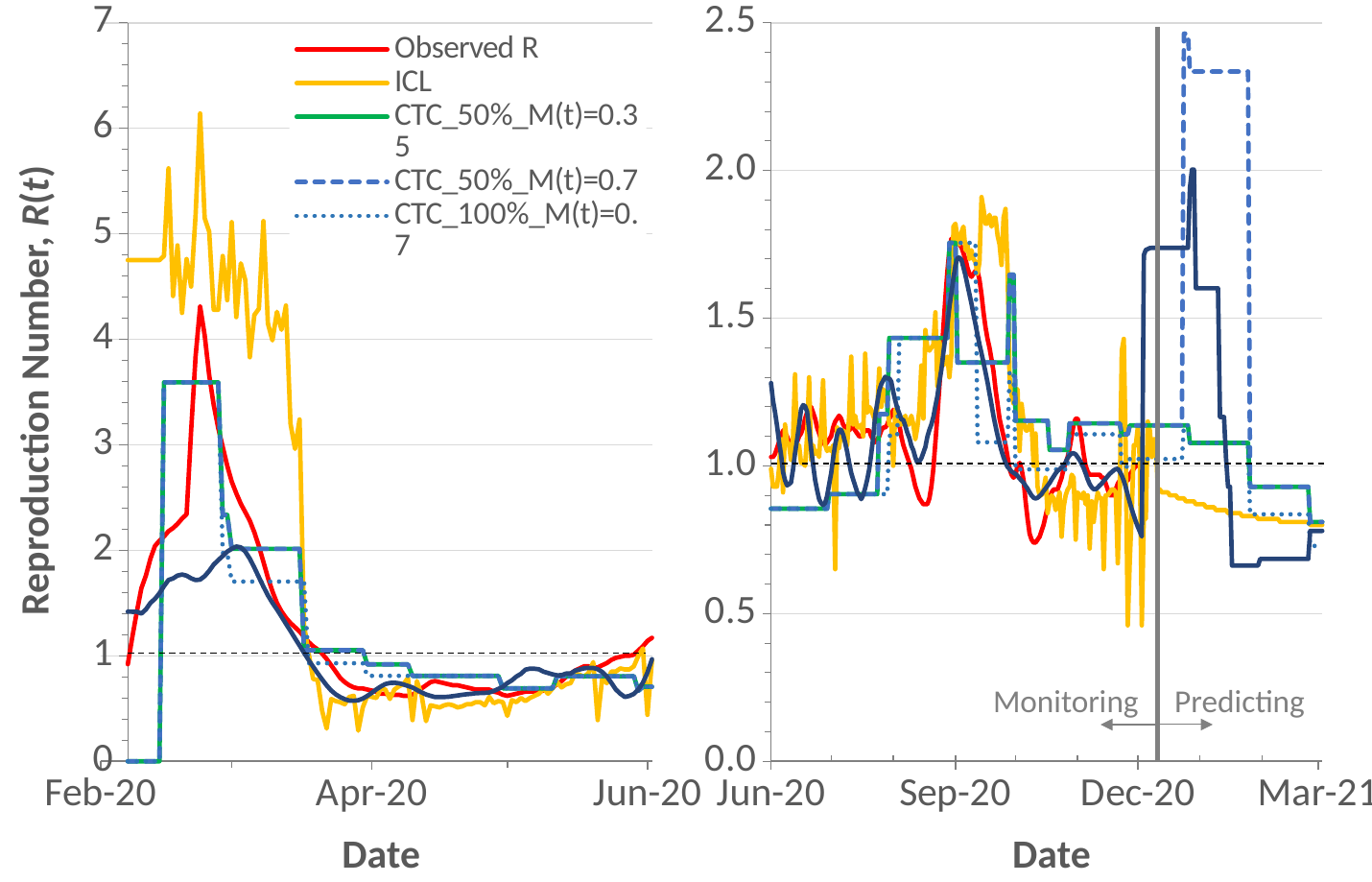

#### Slide 3
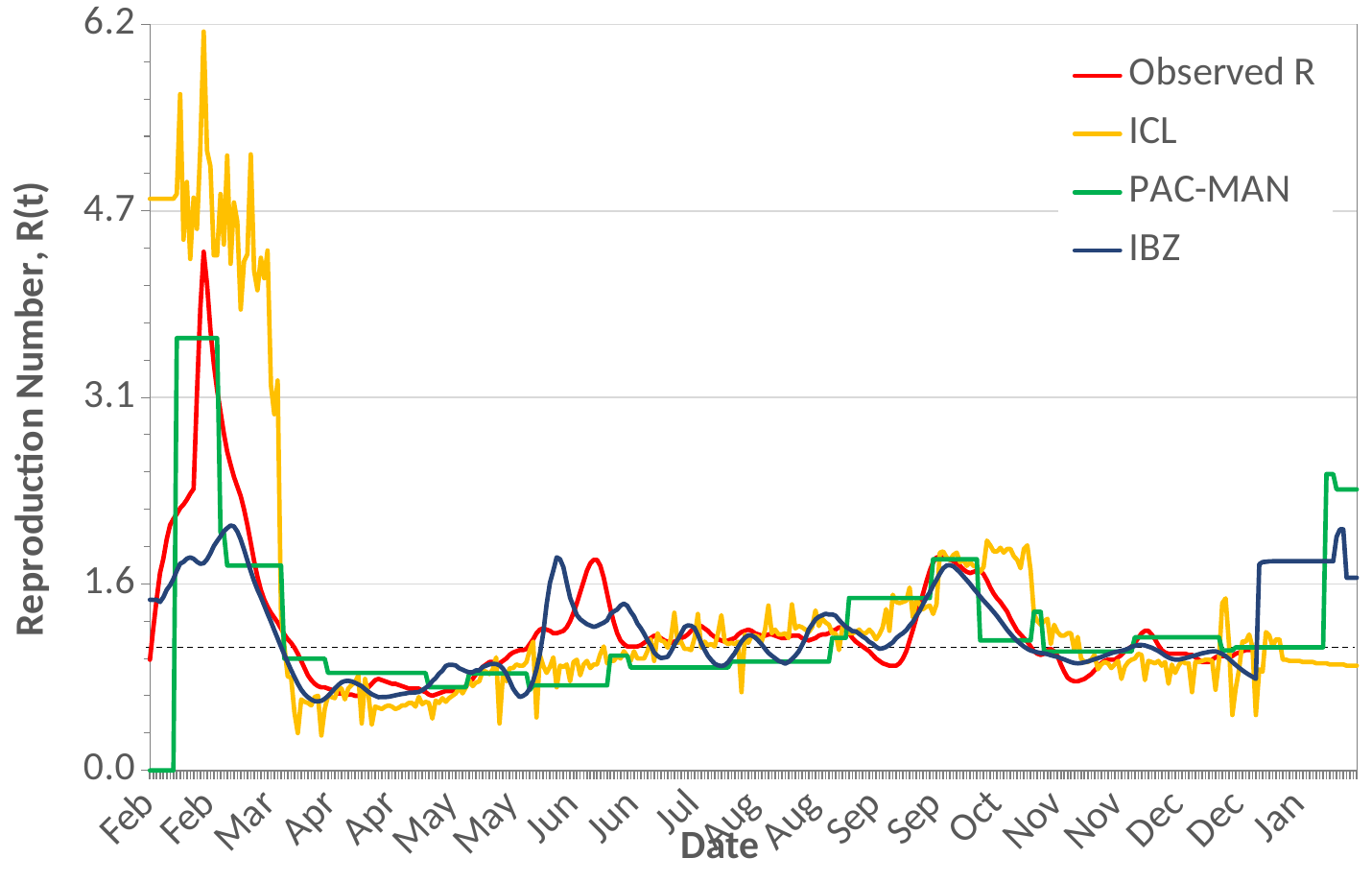

#### Slide 4
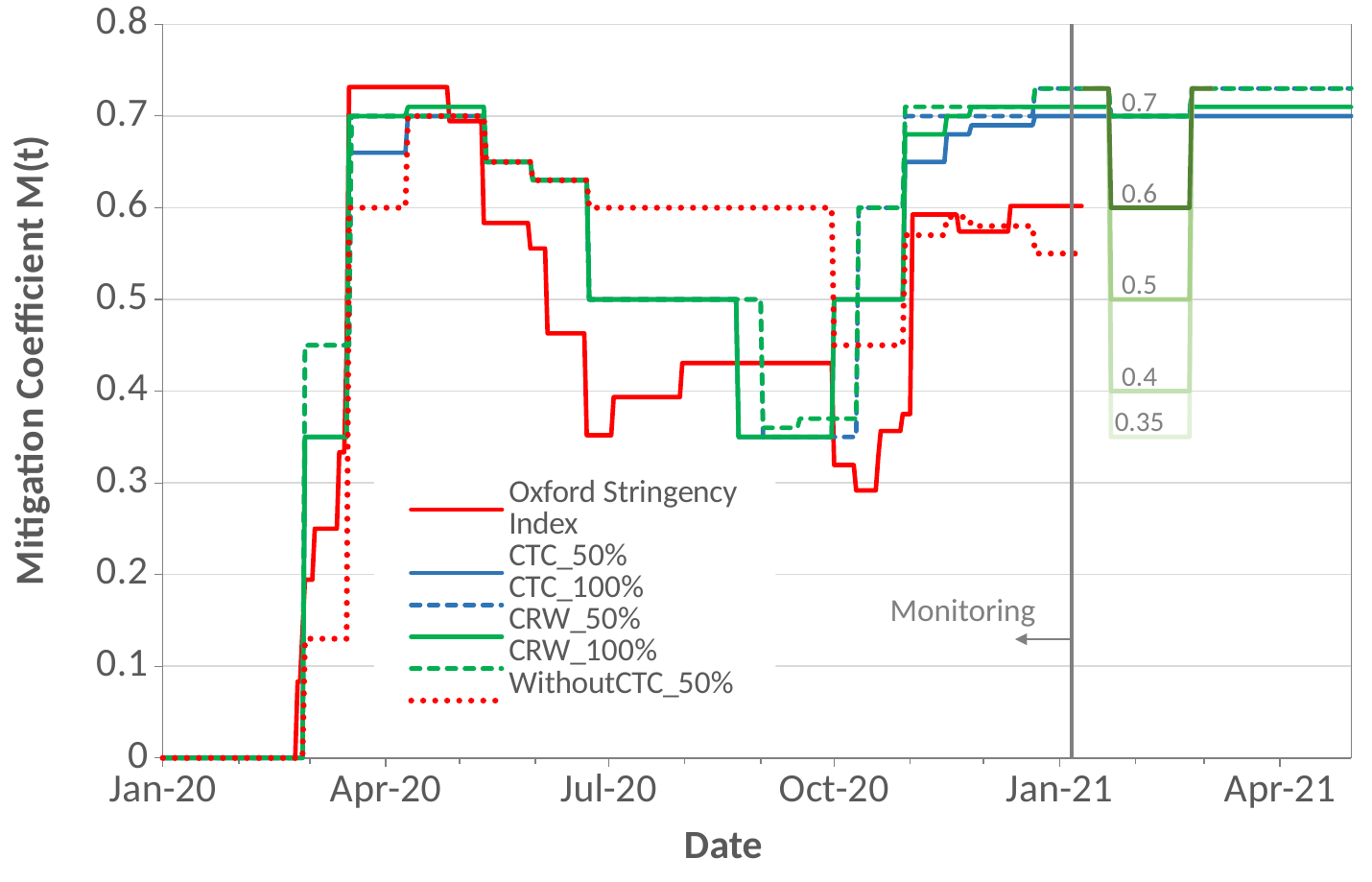

#### Slide 5
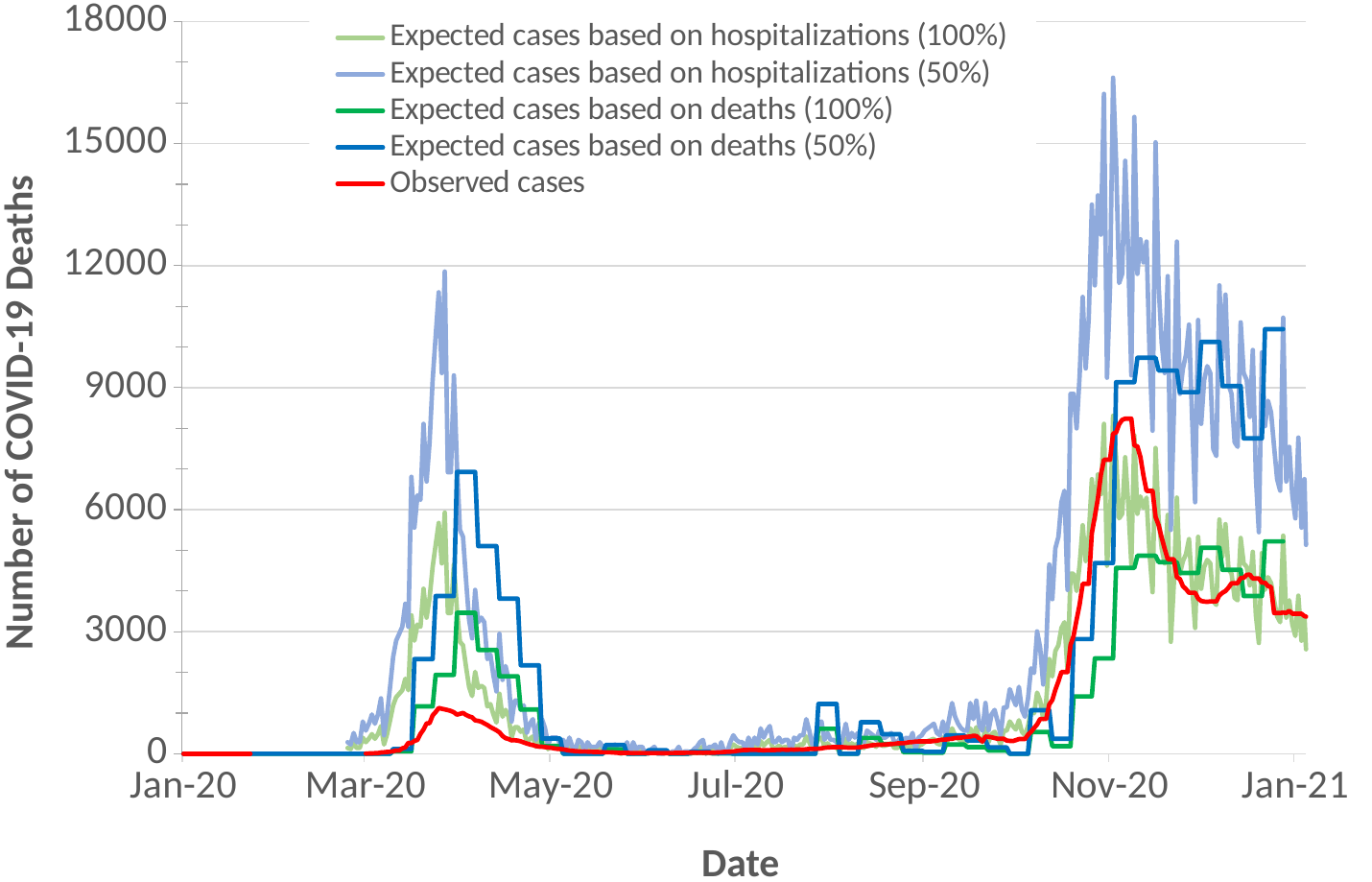

#### Slide 6
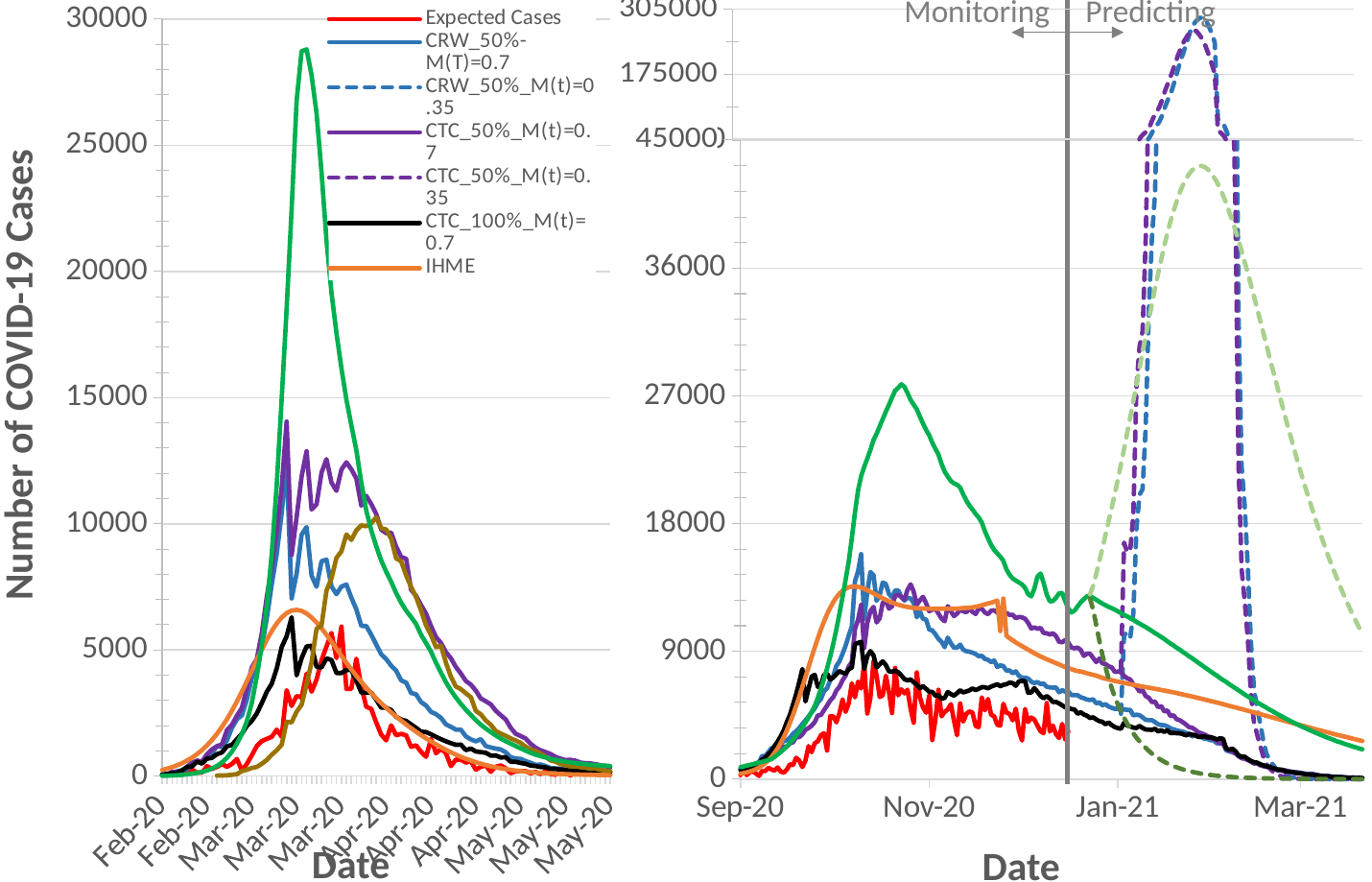

#### Slide 7
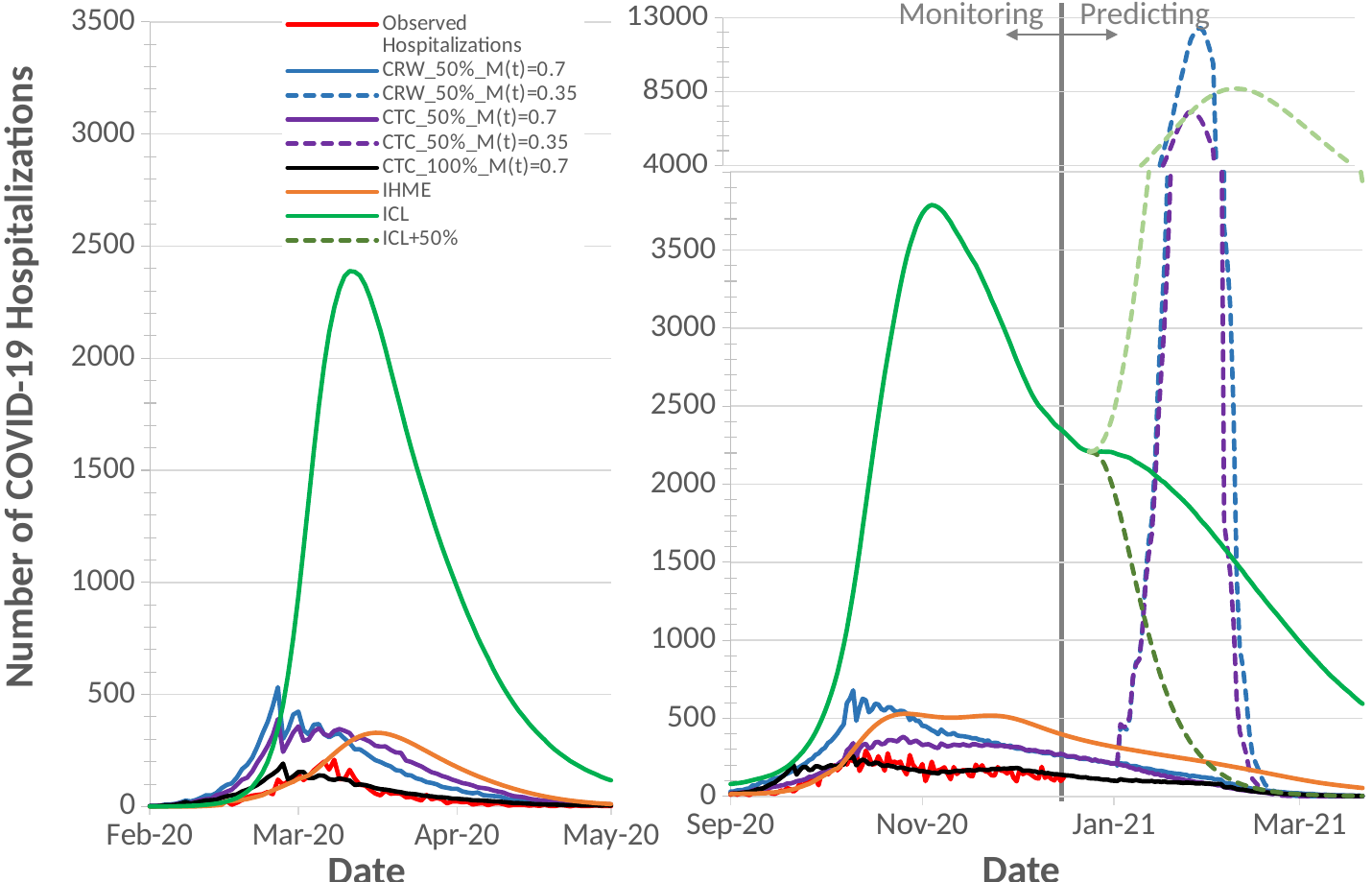

#### Slide 8
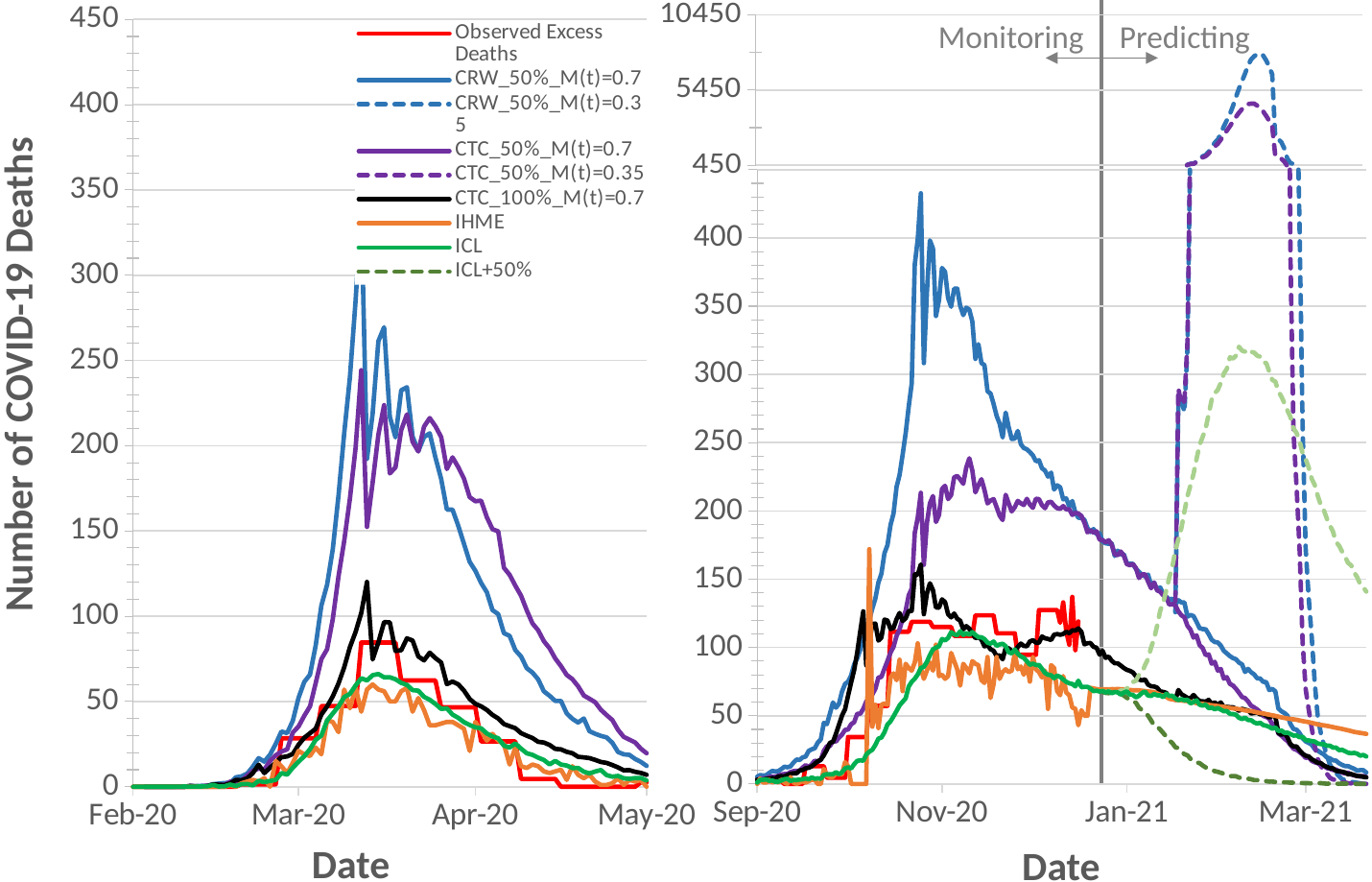

#### Slide 9
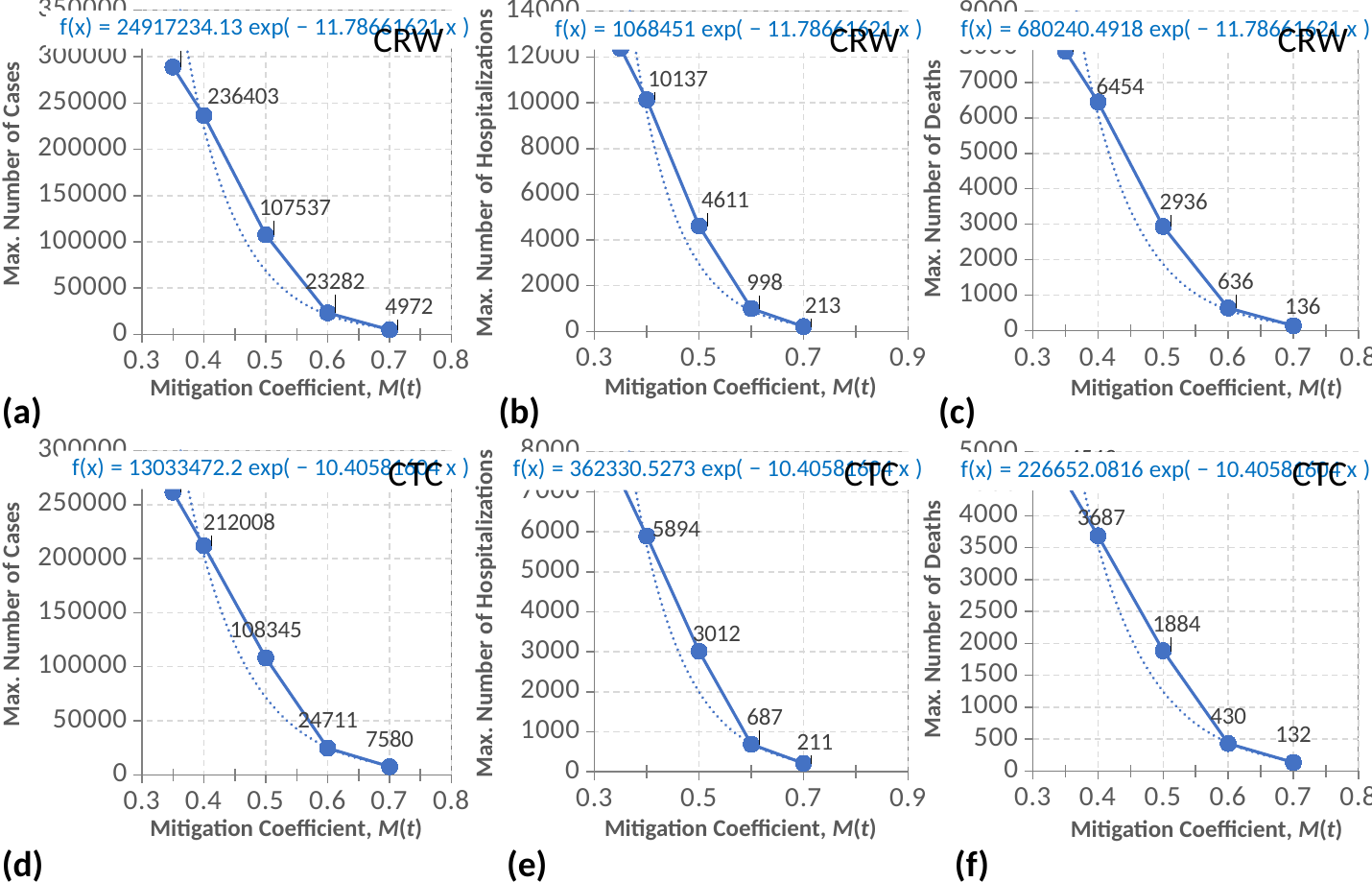

#### Slide 10
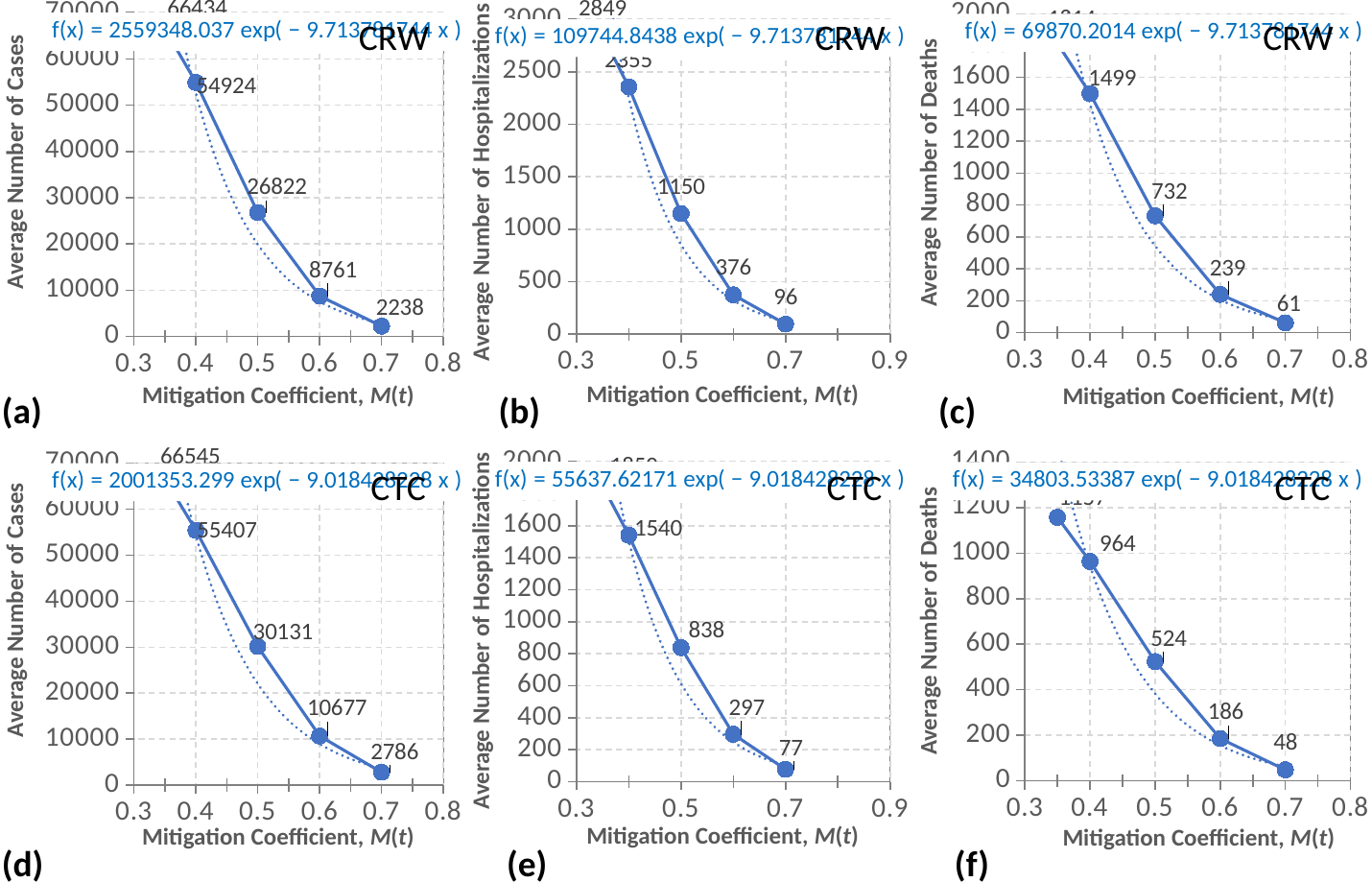
